# Anatomy of digital contact tracing: role of age, transmission setting, adoption and case detection

**DOI:** 10.1101/2020.07.22.20158352

**Authors:** Jesús A. Moreno López, Beatriz Arregui García, Piotr Bentkowski, Livio Bioglio, Francesco Pinotti, Pierre-Yves Boëlle, Alain Barrat, Vittoria Colizza, Chiara Poletto

## Abstract

The efficacy of digital contact tracing against COVID-19 epidemic is debated: smartphone penetration is limited in many countries, non-uniform across age groups, with low coverage among elderly, the most vulnerable to SARS-CoV-2. We developed an agent-based model to precise the impact of digital contact tracing and household isolation on COVID-19 transmission. The model, calibrated on French population, integrates demographic, contact-survey and epidemiological information to describe the risk factors for exposure and transmission of COVID-19. We explored realistic levels of case detection, app adoption, population immunity and transmissibility. Assuming a reproductive ratio *R* = 2.6 and 50% detection of clinical cases, a ~20% app adoption reduces peak incidence by ~35%. With *R* = 1.7, >30% app adoption lowers the epidemic to manageable levels. Higher coverage among adults, playing a central role in COVID-19 transmission, yields an indirect benefit for elderly. These results may inform the inclusion of digital contact tracing within a COVID-19 response plan.

## Introduction

Intervention measures aiming at preventing transmission have been key to control the first wave of the COVID-19 pandemic. Many countries have adopted lockdown and strong social distancing during periods of intense epidemic activity to suppress the epidemic and reduce hospital occupancy below saturation levels (*1, 2*). Due to their huge economic and societal costs these interventions can only be implemented for a limited amount of time. The building of population immunity has been slow (*3–5*), so that new waves are possible after temporary lockdowns and lifting of restrictions. Sustainable strategies are required to maintain the epidemic under control while enabling the close-to-normal functioning of the society. Widespread testing, case finding and isolation, contact-tracing, use of face masks and enhanced hygiene are believed to be crucial components of these strategies.

Contact-tracing aims to avoid transmission by isolating at an early stage only those individuals who are infectious or potentially infectious, in order to minimize the societal costs associated to isolation. Considerable resources are therefore directed at improving surveillance capacities to allow efficient and rapid investigation and isolation of cases and their contacts. To enhance tracing capacities, the use of digital technologies has been proposed, leveraging the wide-spread use of smartphones. Therefore, proximity-sensing applications have been designed and made available – e.g. in Australia, France, Germany, Iceland, Italy, Switzerland – to automatically trace contacts, notify users about potential exposure to COVID-19 and invite them to isolate.

Empirical studies of the impact of these digital applications are however limited (*6–8*), and the utility of this intervention is debated. Some built-in features make it more efficient than manual contact tracing: it is automated, reducing the burden of manual contact tracing and limiting recall bias; it is faster, as information can be transmitted in real time. However, coverage is uneven. In particular, most children and elderly do not own a smartphone or are less familiar with digital technologies. The overall adoption of the app among smartphone owners will also be a limiting factor, as well as the fraction of cases actually triggering the alert to the contacts and the adherence to isolation of the app adopters who receive an alert.

These variables must be gauged in light of the risk factors for exposure and transmission driving the COVID-19 epidemic. First, individuals of different age contribute differently to the transmission dynamics of COVID-19. Younger individuals tend to have more contacts than adults or the elderly. On the other hand, a marked feature of COVID-19 is the strong age imbalance among cases (*9–13*), that may be explained by both a reduced susceptibility (*9, 10*) and an increased rate of subclinical infections in children compared to adults (*10, 11, 13*). As subclinical cases are harder to detect, this implies that identification of cases and of their contacts may be dependent on age. Second, SARS-CoV-2 transmission risk varies substantially by setting. Transmissions were registered predominantly in households, in specific workplaces and in the community (linked to shopping centers, meals, parties, sport classes, etc.) (*14, 15*). This is due, at least in part, to the higher risk of contagion of crowded and indoor environments (*14–16*). Notably, contacts occurring in the community are also the ones more affected by recall biases, thus more difficult to trace with manual contact tracing.

Several modelling studies have quantified the impact of contact tracing (*17–26*), with some of them addressing specific aspects of digital contact tracing (*18–23*). Still the interplay between age and setting heterogeneity in determining the efficacy of this intervention is largely unexplored. Here we provide a systematic exploration of the different variables at play. We considered France as a case study and integrated different sources of data to realistically describe the French population, in terms of its demography and social contact behavior. We accounted for the dynamics of contacts according to age and setting, and for the setting-specific risk of transmission. We used COVID-19 epidemiological characteristics for parametrization. We then modelled case detection and quarantining, isolation of their household contacts and digital contact tracing, under different hypotheses of potential reduction in transmissibility due to other effects (e.g. face-masks and increased hygiene). We quantified the impact of digital contact tracing on the whole population and on different population groups and settings, as a function of several variables such as the rate of app adoption, the probability of detection of clinical and subclinical cases, population immunity, compliance to isolation and transmission potential. Our results provide quantitative information regarding the impact of digital contact tracing within a broader response plan.

## Results

### Dynamic multi-setting contact network

We modelled the French population integrating available demographic and social-contact data. We collected population statistics on age, household size and composition (Figure 1 A, B), workplace and school size, smartphone penetration (Figure 1 E), and commuting fluxes. Then, by following standard approaches in the literature (*27, 28*) individuals were created in-silico with a given age and assigned to a municipality, a household, and a workplace/school according to the statistics. Smartphones were assigned to individuals depending on their age according to available statistics on French users (Figure 1 E) (*29*). Overall smartphone penetration was 64%, that represents the upper bound limit of app adoption in the population – reached when 100% of individuals owning a smartphone download the app. This synthetic population reproduced the location statistics of individuals in different settings, yielding the basis of a multi-setting network of daily face-to-face contacts in household, school, workplace, community and transport (Figure 1 H) (*30–32*). We parametrized the network from a social contact survey providing information on contacts by age and setting (*33*) (Figure 1 C, D). As contacts may occur repeatedly, we associated an activation rate to each contact and sampled each day contacts based on their activation rate (Figure 1 G). We imposed that 35% of the contacts registered during one day occur with daily frequency, as found in (*33*). Figure 1 F and I show that the features of the resulting daily contact network matched the data: the distribution of the number of contacts was right-skewed as the empirical one reported in (*33*) and the contact matrix showed age assortativity and the characteristic parent-children (off-diagonal) contact pattern. As a case study we restricted our study to a municipality with a population size of ~100,000 individuals (see Material and Methods and Supplementary Material for additional details).

**Figure 1.**
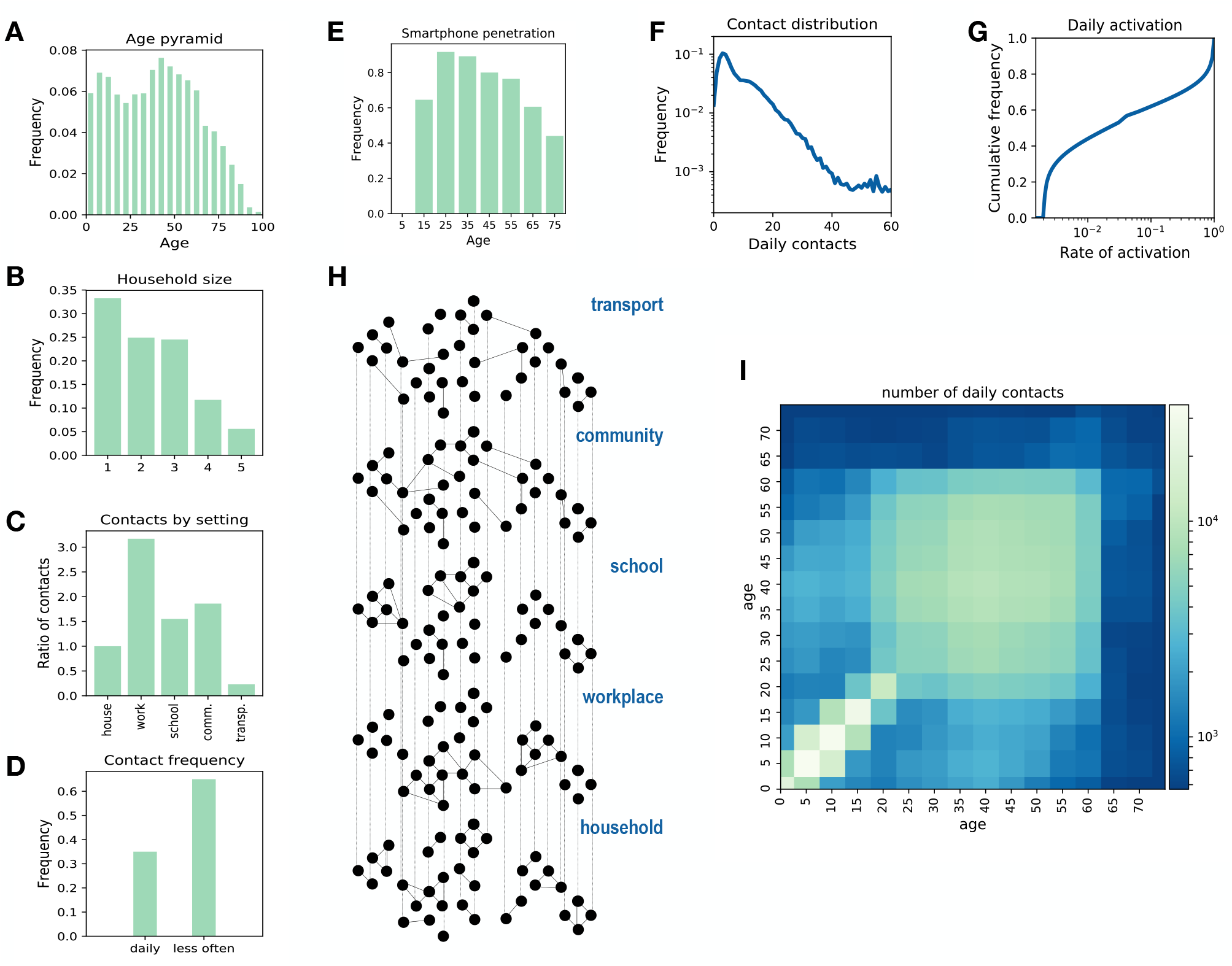
Synthetic population. A-E. Key statistics used as input for the synthetic population reconstruction. **A** Age pyramid for France (source INSEE). **B** Household size (source INSEE). **C** Ratio of contacts by setting with respect to household contacts (*33*). **D** Fraction of contacts occurring each day or less frequently (*33*). **E** Smartphone penetration by age. The overall average adoption in the population is 64% (*29*). **F** Distribution of the number of daily contacts in the model. **G** Cumulative distribution of the activation rate associated to the contacts in the model, calibrated in order to be consistent with the information of panel D. **H** Sketch of the construction of the contact network: contacts among individuals were represented as a multi-layer dynamical network, where each layer includes contacts occurring in a specific setting. **I** Age contact matrix computed from the contact network model.

### COVID-19 epidemic dynamics

We modelled coronavirus transmission and outcome as shown in Figure 2 A, B. Individuals could be susceptible, *S*, exposed, *E*, pre-symptomatic preceding subclinical infection, *I*_*p,sc*_, pre-symptomatic preceding clinical infection, *I*_*p,c*_, subclinically infectious, *I*_*sc*_, clinically infectious, *I*_*c*_, and recovered, *R*. Subclinical cases had symptoms that ranged from no symptoms to mild and continued their normal activity throughout the infectious period.

**Figure 2.**
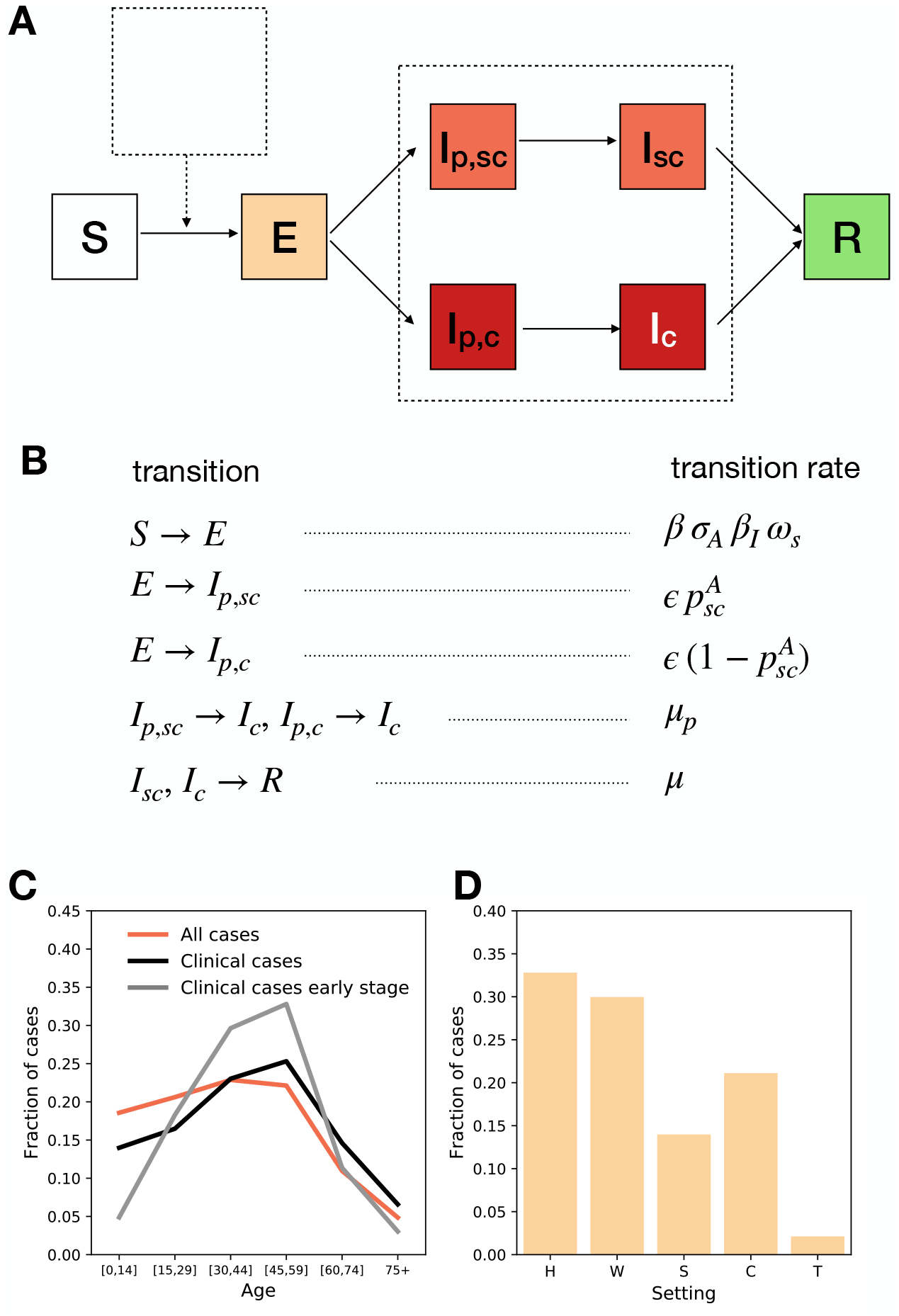
Modelling COVID-19 epidemic. **A, B** Compartmental model summarizing the epidemic states and transitions between states. Parameters and their values are reported in Table 1. **C** Cases by age for an uncontrolled epidemic. We show all cases (clinical and subclinical) in red and clinical cases in black. The grey line shows the clinical cases in the early stage of the epidemic (here defined as the first 30 days), with less cases among children than in later stages. **D** Transmission by setting (H, W, S, C, T stand respectively for household, workplace, school, community, transport). The simulations were done with *β* = 0.25 corresponding to *R*_*0*_ = 3.1. Additional aspects of the outbreak are reported in the Supplementary Material.

Clinical cases had moderate to critical symptoms and stayed at home after the onset of symptoms (*11, 13*) – we did not consider hospitalization. Individuals in compartments *I*_*p,sc*_, *I*_*p,c*_, *I*_*sc*_, *I*_*c*_ transmitted the infection, with subclinical individuals characterized by a lower risk of transmission than clinical ones (see Material and Methods). We accounted for the heterogeneous susceptibility and clinical manifestation by age as parametrized from (*9, 13*) (Table 1). In order to parametrize the infection’s natural history, we combined evidence from epidemiological and viral shedding studies. We used 5.2 days for the incubation period (*34*), 2.3 days for the average length of the pre-symptomatic phase (*35*), and 7 days on average for the infectivity period after symptoms’ onset (*35*).

**Table 1.**
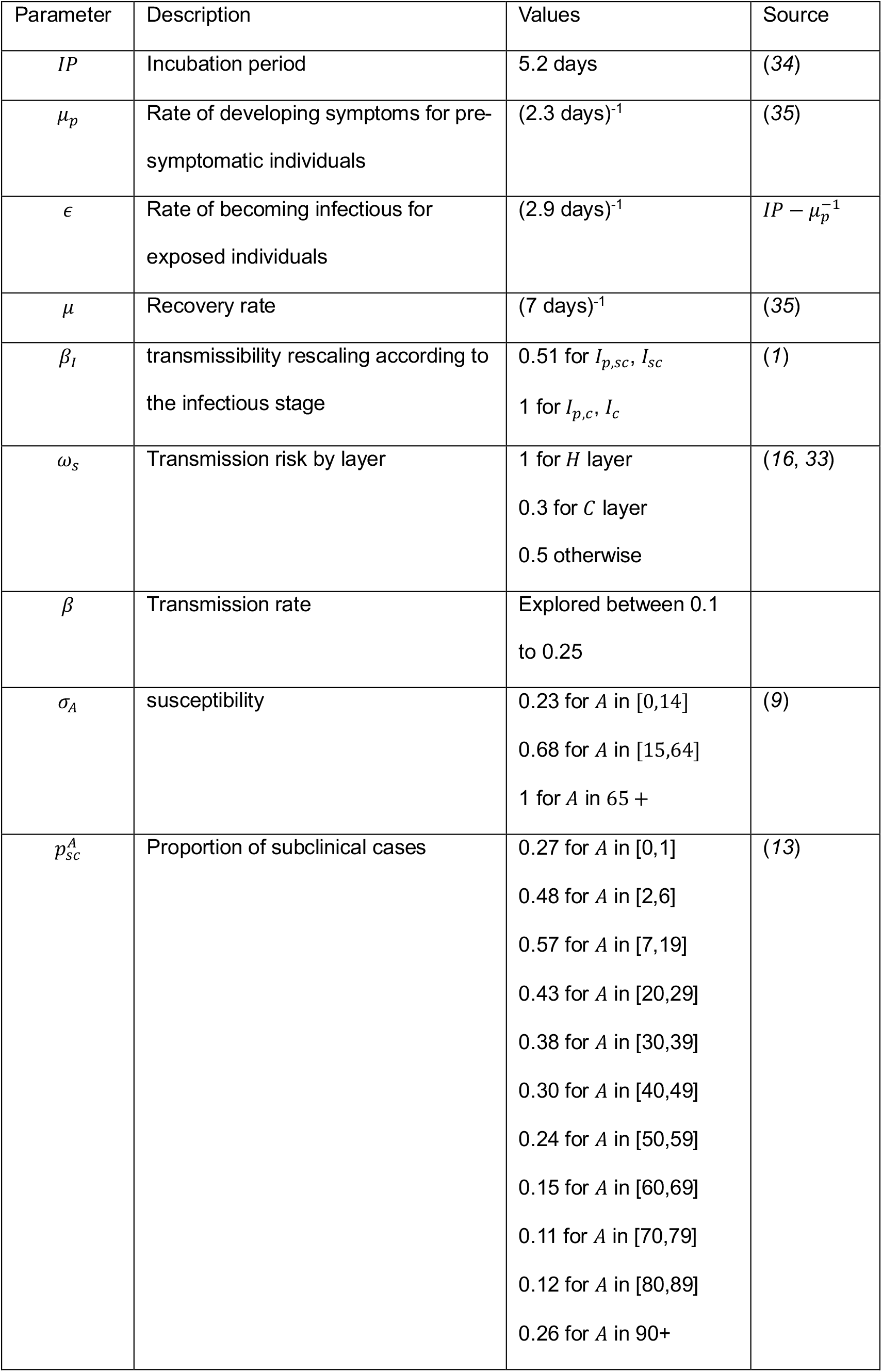
Compartmental model parameters and their values.

| Parameter | Description | Values | Source |
| --- | --- | --- | --- |
| $IP$ | Incubation period | 5.2 days | (34) |
| $\mu_p$ | Rate of developing symptoms for pre-symptomatic individuals | $(2.3 \text{ days})^{-1}$ | (35) |
| $\epsilon$ | Rate of becoming infectious for exposed individuals | $(2.9 \text{ days})^{-1}$ | $IP - \mu_p^{-1}$ |
| $\mu$ | Recovery rate | $(7 \text{ days})^{-1}$ | (35) |
| $\beta_I$ | transmissibility rescaling according to the infectious stage | 0.51 for $I_{p,sc}, I_{sc}$<br>1 for $I_{p,c}, I_c$ | (1) |
| $\omega_s$ | Transmission risk by layer | 1 for $H$ layer<br>0.3 for $C$ layer<br>0.5 otherwise | (16, 33) |
| $\beta$ | Transmission rate | Explored between 0.1 to 0.25 | |
| $\sigma_A$ | susceptibility | 0.23 for $A$ in $[0,14]$<br>0.68 for $A$ in $[15,64]$<br>1 for $A$ in $65 +$ | (9) |
| $p_{sc}^A$ | Proportion of subclinical cases | 0.27 for $A$ in $[0,1]$<br>0.48 for $A$ in $[2,6]$<br>0.57 for $A$ in $[7,19]$<br>0.43 for $A$ in $[20,29]$<br>0.38 for $A$ in $[30,39]$<br>0.30 for $A$ in $[40,49]$<br>0.24 for $A$ in $[50,59]$<br>0.15 for $A$ in $[60,69]$<br>0.11 for $A$ in $[70,79]$<br>0.12 for $A$ in $[80,89]$<br>0.26 for $A$ in $90+$ | (13) |

We first simulated an uncontrolled epidemic assuming transmission levels corresponding to *R*_0_ = 3.1, within the range of values estimated for COVID-19 in France at the early stage of the pandemic (*1, 36*). The generation time resulting from our model and parameters had mean value of 6.0 days (95% CI [2,17]), in agreement with epidemiological estimates (*11, 35, 37*). Figure 2 C and D show the repartition of cases among age groups and settings at the early stage and during the whole course of the epidemic. Age-specific infection probability was higher among young adults, while clinical infections were shifted towards older population with respect to the overall (clinical and subclinical) cases, as noted in previous observational and modelling works (*10*). The age profile changed in time with children infected later as the epidemic unfolded (*10, 38*). Transmissions occurred predominantly in household and workplaces followed by the community setting (*14*).

### Contact tracing

We quantified the impact of combined household isolation and digital contact tracing considering the possible scenario of a new epidemic wave emerging after the release of strict lockdown measures in the country. We thus assumed some level of immunity to the virus – exploring a range from 0 to 15% of the population. We considered interventions based on the use of digital contact tracing, coupled with testing and isolation of clinical cases and households. 50% of individuals with clinical symptoms were assumed to get tested after consulting a doctor and to isolate if positive. Higher and lower percentages were also considered.

Case tracing was assumed to start when a case with clinical symptoms tested positive and was isolated, with an average delay of ~1 day. Household members were also invited to isolate – we assumed that 90% of them accepted to isolate and that their isolation occurred at the same time as the detected case. If the index case had the app installed, the contacts he/she registered in the previous *D* = 7 days were notified and could decide to isolate with a compliance probability of 90% – lower values of compliance were also explored. Note that only contacts occurring between individuals who both use the app can be registered, so only app adopters can be notified. We explored several levels of app adoption in the population. In addition to the detection of clinical cases, we assumed that a proportion of subclinical cases was also identified. These may be cases with very mild, unspecific symptoms who decided to get tested as part of vulnerable groups (i.e. co-morbidity) or because highly exposed to the infection (health care professionals). We hypothesized this proportion to be small in the baseline scenario (5%), and we later varied it up to 45%. Isolated individuals resumed normal daily life if infection was not confirmed. We took 7 days as the time needed for being confirmed negative because multiple tests and some delay since the exposure are needed for a negative result to be reliable. Infected individuals got out of quarantine after 14 days unless they still have clinical symptoms after the time is passed. They may, however, decide to drop out from isolation each day with a probability of 2% if they don’t have symptoms (*21, 26*).

Figure 3 summarizes the effect of the interventions. We compared the uncontrolled scenario (*R* = *R*_0_ = 3.1) with scenarios where the transmissibility is reduced due to the adoption of barrier measures (*R* down to 1.5). We also assumed 10% of the population to be immune to the infection (*36*). Panels A-C shows the results for *R* = 2.6 and *R* =1.7. With *R* = 2.6 (Figure 3 A, C), the relative reduction of peak incidence due to household isolation only would be 27%. The inclusion of digital contact tracing would increase the relative reduction to 35% with ~20% app adoption, and to 66% with ~60% app adoption – i.e., 90% of individuals owning a smartphone use the app. This corresponds to an additional mitigation effect ranging from 30% to 144% provided by contact tracing compared to household isolation only. With *R* =1.7 (Figure 3 B, C), we find that ~20% app adoption would reduce the peak incidence by 45% (additional mitigation effect of 25%), while the reduction would reach 89% in a scenario of ~60% app adoption (additional mitigation effect of 147%). According to the projections in Ref.(*1*), intensive care units occupation would remain below the saturation level with incidence below 0.4 /1000 hab. In the scenario with *R* =1.7, this would be reached with app adoption greater than ~30% (grey dashed line in Figure 3 B). Stronger reductions could be obtained with more efficient detection of clinical cases (Figure 3 E, H, obtained with *R* = 2.6) and of subclinical ones (Figure 3 L, *R* = 2.6). The relative reduction in peak incidence produced by ~20% app adoption would be 47% with an 80% detection rate of clinical cases, compared to the 35% relative reduction obtained with 50% detection rate. Results show similar trends across different levels of population immunity, with higher relative impacts predicted for low immunity (Figure 3 F, I, *R* = 2.6). Compliance to isolation of household contacts had an appreciable effect at low app adoption (Figure 3 K, *R* = 2.6). 50% compliance would reduce peak incidence of 19%, compared to 27% reduction for 90% compliance, in the case of household isolation only. Compliance of notified contacts to isolation, instead, has a larger effect on peak incidence only when app adoption is high, as expected. For example, if the app was adopted by 60% of individuals the reduction in peak incidence would pass from 55% to 66% if compliance changed from 50% to 90% (Figure 3 J, *R* = 2.6).

**Figure 3.**
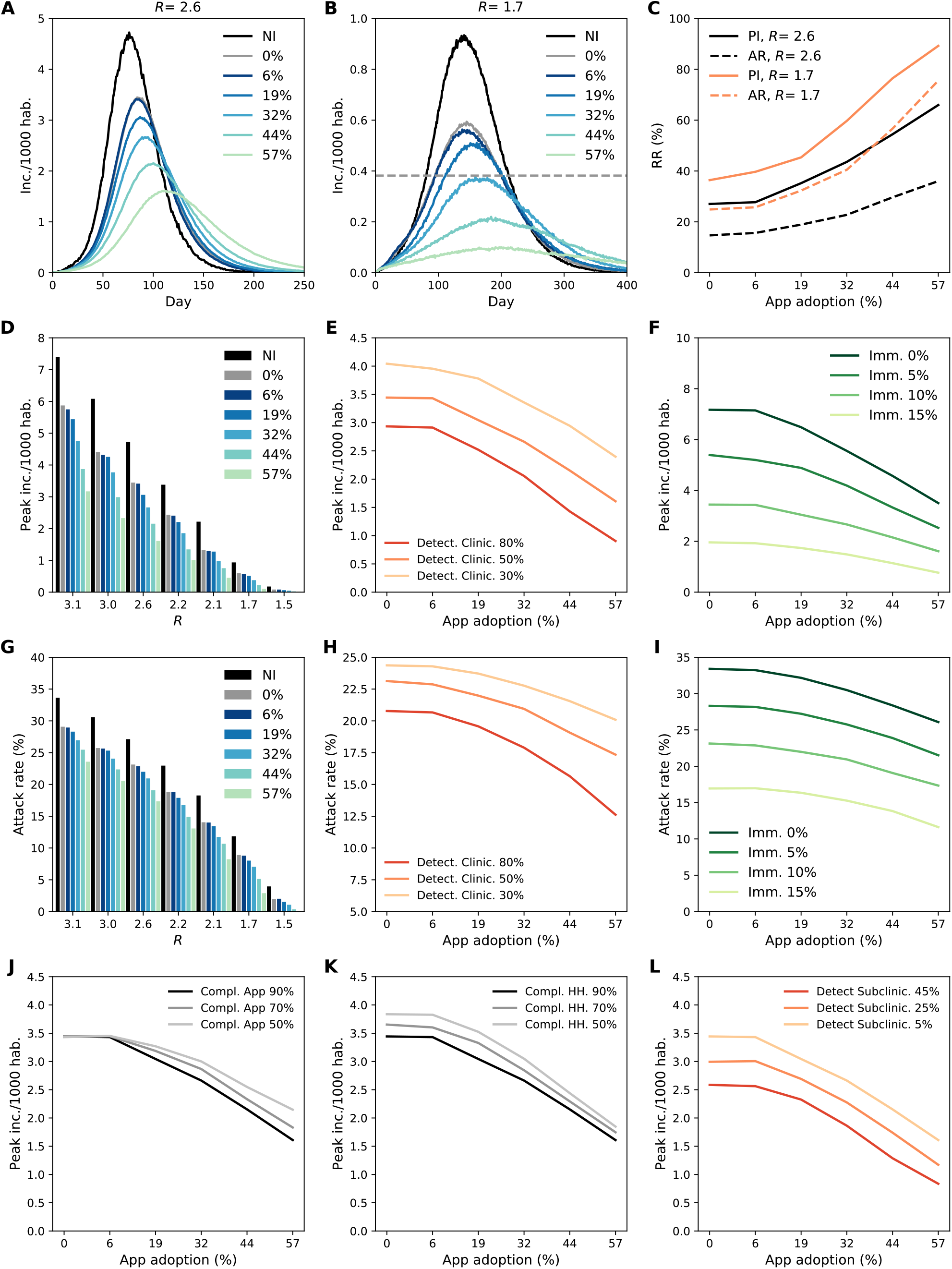
Impact of digital contact tracing and household isolation on the epidemic. A, B. Incidence (clinical cases) according to app adoption for *R* = 2.6 and *R* =1.7, respectively. The black curve shows the scenario with no intervention (NI). Other curves correspond to app adoption levels ranging from 0% (household isolation only) to 57% (90% of smartphone users). Incidence threshold level corresponding to ICU saturation is showed as a dashed grey line in panel B. **C** Relative reduction (*RR*) in attack rate (AR) and peak incidence (PI) as a function of the app adoption for the scenarii shown in A and B. *RR* is computed as 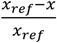, where *x* is either PI or AR and *x* is the value of the quantity with no intervention. Attack rate is computed as cumulative incidence discounting initial immunity (10%). **D, G** Peak incidence and attack rate according to reproduction ratio *R* and app adoption. **E, H** Peak incidence and attack rate according to app adoption and percentage of clinical cases detected. **F, I** Peak incidence and attack rate according to app adoption and initial immunity. **J** Peak incidence according to app adoption and compliance to isolation of contacts notified by the app. **K** Peak incidence according to app adoption and compliance to isolation of household contacts. **L** Peak incidence according to app adoption and percentage of subclinical cases detected. Except as otherwise indicated, parameters values were: initial immunity 10%, clinical case detection 50%, subclinical case detection 5%, compliance to isolation of contacts notified by the app 90%, compliance to isolation of household contacts 90%, *R* = 2.6.

We analyzed the simulation outputs to characterize index cases and their contacts and relate this to the reduction in number of cases by age and setting. We found that adults represented the majority of index cases (Figure 4 D), while their household contacts were mostly children. The app registered mostly contacts with adults, and the tracked contacts were occurring predominantly in workplaces and in the community (Figure 4 A). This results in a heterogenous reduction in transmission (*TRR*) by setting and age group. Household isolation reduced transmission in all settings, with the smallest effect in workplaces (Figure 4 B). Digital contact tracing has instead a high *TRR* at work, in the community and in transports (Figure 4 C). Household isolation reached mostly children (< 15 years old) and the elderly (especially the 75+ group) with the smallest effect in the 15-59 years old (Figure 4 E). Adopting digital contact tracing led to an increased TRR with age, even among the oldest age range (Figure 4 F). This result shows the indirect effect of digital tracing: due to the central role of adults in the transmission of SARS-CoV-2 towards all age-groups, avoiding adult infections led to less transmission to the elderly. We also tested the case in which elderly people (70+) owning a smartphone did not install the app at all, assuming they are less familiar with digital technologies, and we found no appreciable effect. These results and additional details are provided in the Supplementary Material.

**Figure 4.**
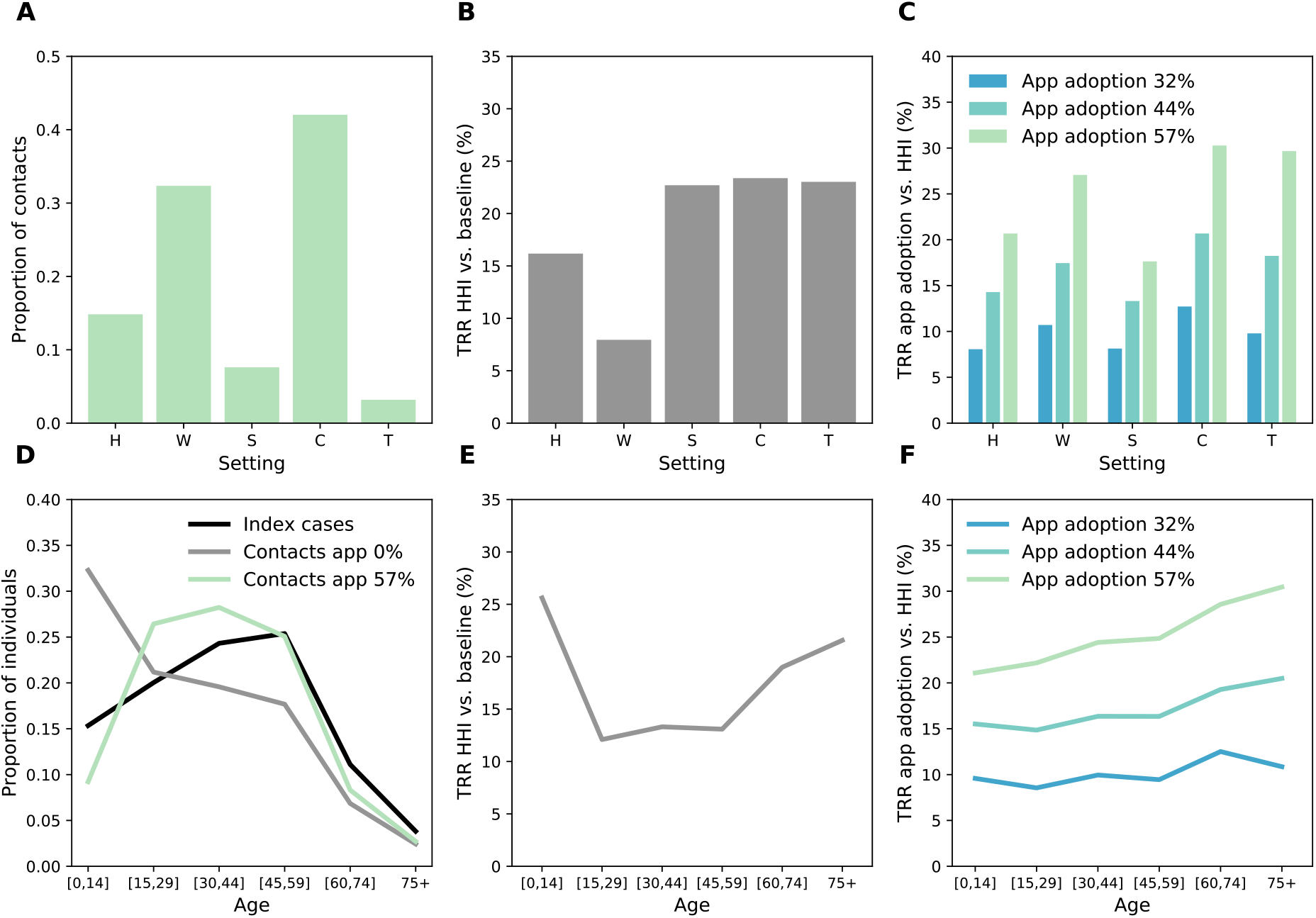
Effect of digital contact tracing and household isolation by age and setting. A. Repartition among the different settings of the contacts detected by contact tracing (57% app adoption). **B** Relative reduction in transmission (*TRR*) by setting obtained with household isolation. The relative reduction in transmission is here defined as 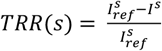, where *I* is the total number of clinical and subclinical cases infected in setting *s*, in the given intervention scenario considered (here household isolation) and 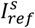 is the same quantity in the reference scenario (here the scenario with no intervention). **C** *TRR* obtained with digital contact tracing with respect to household isolation only, for three values of app adoption. **D** Repartition among the different age groups of the index cases and of the detected contacts, in a scenario with household isolation only, and with the inclusion of digital contact tracing (57% app adoption). The repartition of index cases is very similar in the two scenarios, thus only the one with household isolation is shown for the sake of clarity. **E** *TRR* by age group of the infected as obtained with household isolation only. **F** *TRR* of digital contact tracing with respect to household isolation only. We assume *R* = 2.6, immunity 10% and probability of detection 50%.

### Traced and Isolated individuals

Feasibility of contact tracing depends on the number of traced contacts who require assistance and virological tests. In a scenario with high detection rate (80%), we found that for each detected case 1.5 contacts were identified on average through household isolation but up to 7.5 with app adoption at 57%, for *R* = 2.6 and 10 for *R* =1.7 (Figure 5 D). This number was however subject to fluctuations (Figure 5 A). Overall, the maximal fraction of the population quarantined at any given time was ~50 per 1000 habitants in a scenario with *R* = 2.6, and was between ~1 and ~4.5 per 1000 habitants when *R* =1.7 (Figure 5 B and E). The latter case corresponded to the situation in which high levels of app adoption were able to strongly reduce spreading, thus the proportion of isolated individuals declined in time, signaling the success of quarantining in preventing the propagation of the infection. A total of 30 per 1000 habitants were isolated in a scenario with *R* =1.7, assuming high app adoption. At *R* = 2.6, 1030 per 1000 habitants were isolated at the end of the epidemic meaning that certain individuals were isolated more than once. In all scenarios, the increase of app adoption inevitably determined an increase in the proportion of people that were unnecessarily isolated, i.e. of individuals that were not infected but still isolated (Figure 5C, F): this proportion increased from 61% to 84% with the increase of app adoption from 0% to 57% (note that the case of 0% app adoption implies that 61% of individuals who were isolated through household isolation were not infected). These numbers were similar for the two tested values of *R* =1.7 and 2.6 (Figure 5 C and F).

**Figure 5.**
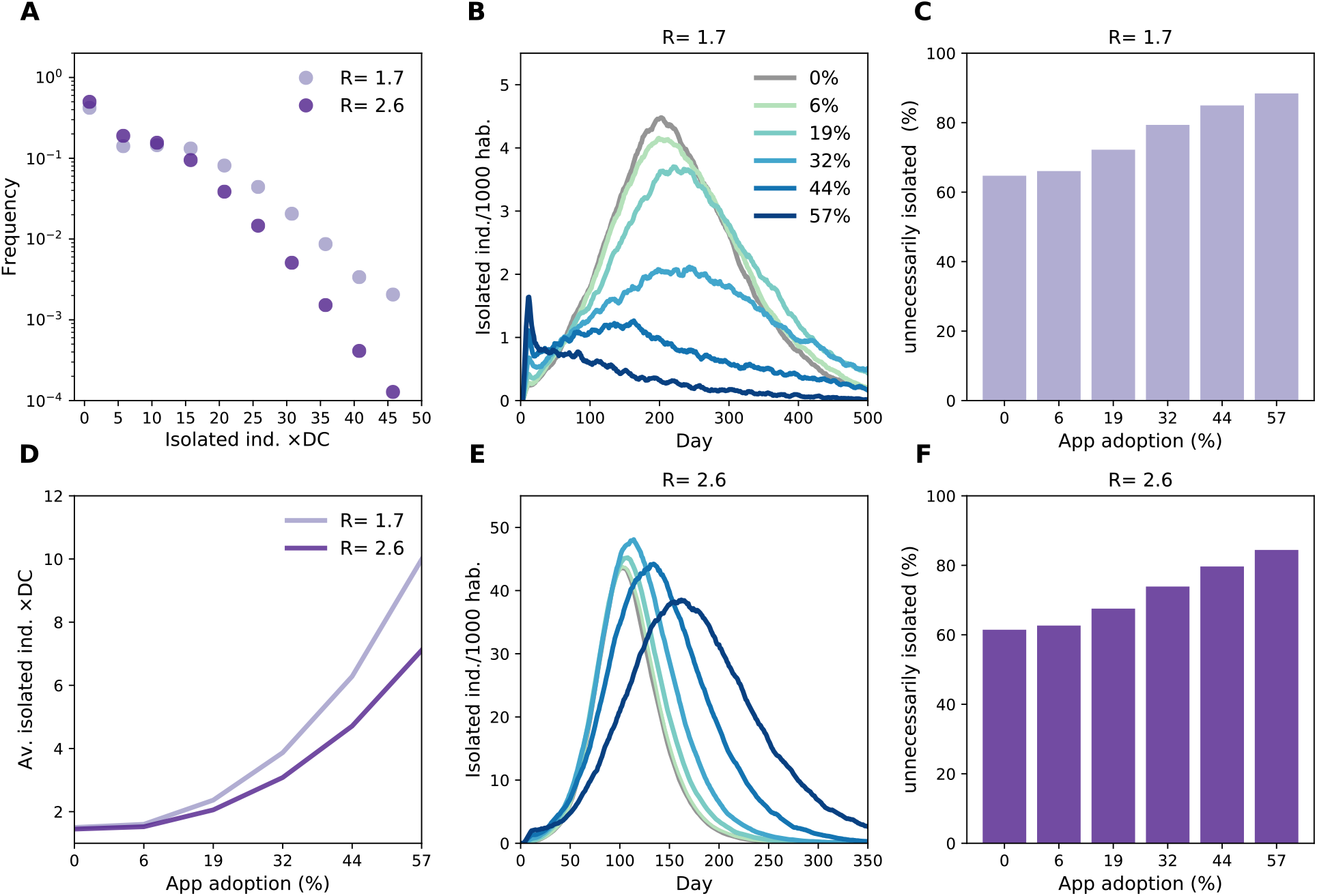
impact of combined digital contact tracing and household isolation on the isolation of individuals. A. Distribution of the number of isolated individuals per detected case (DC) for 57% of app adoption. **D** Average number of isolated individuals per detected case as a function of app adoption. **B, E** Percentage of the population isolated as a function of time for *R* =1.7 (B) *R* = 2.6 (E). **C, F**: Fraction of unnecessary isolated, i.e. fraction of contacts isolated without being positive.

## Discussion

Quantifying the impact of digital contact tracing is essential to envision this strategy within a wider response plan against the COVID-19 epidemic. We modelled this intervention together with household isolation assuming a 50% detection of clinical cases. In a scenario of high transmissibility (*R* = 2.6), we found that household isolation by itself would produce a reduction in peak incidence of 27%, while the inclusion of digital contact tracing could increase this effect by 30% for a reasonably achievable app adoption (~20% of the population), and by 144% for a large-scale app adoption (~60%). At a moderate transmissibility level (*R* =1.7), the app would substantially damp transmission (36% to 89% peak incidence reduction for increasing app adoption), bringing the epidemic to manageable levels if adopted by 32% of the population or more. Importantly, the app-based tracing and household isolation have different effects across settings, the first intervention efficiently preventing transmissions at work that are not well targeted by the second. Moreover, app-based contact tracing also yields a protection for the elderly despite the lower penetration of smartphones in this age category.

Lockdown and social distancing have been effective in reducing transmission in the first epidemic wave in many countries. However, their huge societal and economic costs made their prolonged implementation impossible. Phasing out lockdown occurred at the beginning of the summer in Europe, with high temperatures, increased ventilation and outdoor activities helping reducing the risk of contagion (*16*). The relaxation of almost all restrictive measures, the start of activities in the fall and the cold season accelerated transmission, reaching a point in which strict non-pharmaceutical interventions were again necessary to curb the epidemic increase. At the time of writing, national or local lockdowns were restored in several countries in Europe (*39*). This highlights the need for exit strategies based on sustainable non-pharmaceutical interventions, able to suppress COVID-19 spread while having limited impact on the economy and on individuals’ daily life (*1*).

Many countries have increased their capacity to detect cases and track their contacts. In France, thousands of transmission clusters have been identified and controlled since the end of the first lockdown period (*40*). Under-detection of cases was however estimated over summer, and the system was predicted to deteriorate rapidly for increasing epidemic activity (*41*). The automated tracking of contacts could then provide an important complementary tool. Here we found that digital contact tracing could reduce attack rate and peak incidence, in agreement with previous works (*18, 19, 26*). The impact of the measure would depend on population immunity, thus geographical heterogeneities should be expected, as regions were differentially hit by the first wave of the epidemic (*4*). On the other hand, app adoption as well may be higher in these areas because of risk aversion behavior (*42*). Also, higher participation rates may be expected in dense urban areas to protect from exposure from random encounters (e.g. in public transports).

Under realistic hypotheses, the intervention would not be able alone to bring the epidemic under control in a scenario where transmission is high (*18, 19, 26*), mainly due to the strong role of asymptomatic transmission in fueling the epidemic (*11, 12, 43*). We explored different values of the reproductive number *R*, to effectively account for non-pharmaceutical measures mitigating the epidemic and for the adoption of preventive measures substantially hindering SARS-COV-2 transmission. We found that a reduction of the epidemic to a manageable level would be possible with a moderate *R* (e.g. *R* =1.7 explored here).

Improved case finding is the first step towards a successful contact-tracing intervention. We found that the increase in detection of clinical cases substantially reduced peak incidence and improved the efficacy of contact tracing. Many countries progressively increased testing capacity (*41*) and lifted restrictions on access to testing (*44*). Easy access to testing is essential to detect cases, because of the substantial fraction of subclinical cases and the similarity of COVID-19 clinical presentation to the one of other respiratory infections. In the period from September to November 2020, the French network of virological surveillance run by general practitioners reported that only 22.7% of Acute Respiratory Infections were caused by SARS-CoV2, against 46.5% attributed to rhinovirus (*45*). Given that the majority of cases do not require hospitalization, case detection effectiveness is also influenced by the consultation rate. This has been estimated to be around ~30% with peaks at ~45% by the participatory surveillance platform covidnet.fr (*40, 41*). Increased population awareness is thus essential for the efficient monitoring of the epidemic and its containment through contact tracing.

Little information is available on compliance to isolation. Low compliance to isolation was reported in the UK and in a university campus in the US (*46, 47*). However, this may vary greatly according to cultural, socioeconomic and demographic context. Due to a self-selection bias, individuals who decided to download and use the mobile application may be more akin to follow the recommendation and isolate if they receive a notification. Step-by-step recommendations provided by the app can further help in increasing compliance. Strengthened communication and compensations (such as paid work leave, loss-of-income payments for self-employed professionals, medical school-absence certificates) should be implemented to increase the acceptability of isolation (*48*).

App adoption remains the key factor determining the efficacy of digital contact tracing. Adoption levels were initially low (<5%) in many countries (e.g. Italy, France), increasing later as the second wave was rising, likely due to increased concerns of the population. As of November 2020, 17% and 13% of the population had downloaded the app in Italy and France, respectively (*49, 50*). Higher levels were observed, e.g., in Australia (6 millions download, 25% of the population) (*51*) and Iceland (~150 thousand, 38%) (*52*). Importantly, official figures may overestimate real adoption levels, since many individuals may download the application without using it. In France this proportion was 60% among university students (*53*). Individuals may be more inclined to use the app if they perceive a direct and immediate benefit from its use. This may be implemented through, e.g., easy access to testing in case they are notified as contacts and assistance by public health professionals. Moreover, even if the application preserves users’ privacy and can be downloaded on a voluntary basis in many countries, increased transparency and ethical debate remain essential to reassure the population about data treatment (*53–55*).

The results presented here are based on an agent-based model that describes age-specific risk factors for exposure and transmission: contact rates, contacts by location, susceptibility to the virus, probability of being detected and rate of app adoption. The interplay between these features has a profound impact on COVID-19 spread and affects the efficacy of household isolation and digital contact tracing. To account for contact heterogeneities we used statistics on population demography, combined with social contact surveys to build a multi-setting contact network, similarly to previous works (*17, 21, 26, 30–32*). The network is also dynamic in time as it captures the repetition of a certain number of contacts (e.g. relationships) and the occurrence of random encounters. Social contact data provide an invaluable information source to study the current COVID-19 outbreak (*1, 36*). Previous projections on the impact of contact tracing rely on a similar approach in some cases (*17, 24, 26*). Other works make use of high resolution data (*18, 19, 22*), that are more reliable than contact surveys, but are restricted to specific settings or population groups. Despite the difference in the data source and approach, the results of these studies are consistent and in agreement with our work on the overall impact of the intervention.

We modelled age-specific epidemiological characteristics based on available knowledge in the literature. Children are less impacted by the COVID-19 epidemic (*9–13*). This may be explained by reduced susceptibility and severity, with accumulating evidence that both effects are acting simultaneously (*10*). The strength of these effects is still debated and the infection risk for children should not be minimized. However, these differences have implications for digital contact tracing. Indeed, it is precisely in the group that plays a central role in transmission and where cases are more likely symptomatic (i.e., adults) that the app coverage is already the highest. Our model shows that taken together, these characteristics reinforce the impact of digital tracing and provides indirect protection in the elderly population. This occurs even if no adoption is registered in the elderly population.

Our study is affected by limitations. First, we analyzed the effect of digital contact tracing on COVID-19 incidence in the general population. Crucial information for public health authorities would be to quantify the effect in time of these measures on hospitalizations. This would require to couple our model for COVID-19 transmission in the general population with a model describing disease severity and within-hospital patient trajectories (*17, 21, 26*). Second, the model does not account for transmission in nursing homes. This setting is where the majority of transmissions among elderly occurred. At the same time, however, the response to the COVID-19 epidemic in this setting relies mostly on routine screening of symptoms and frequent testing of residents, together with face masks and strict hygiene rules for visitors. Third, results may be conservative as clustering effects and large fluctuations in the number of contacts per person (*56*) are only partially captured by the model thanks to the repetition of contacts, but effects may be larger in real contact patterns. This also includes crowding events playing an important role in the transmission dynamics (*15*). Overall behaviors obtained with our synthetic network of contacts are however compatible with findings obtained with real contact data (*18*). In a future work the description of temporal and topological properties of contacts in workplaces, schools and community could be improved by using modeling frameworks informed by detailed contact data, that has become available for specific settings (*57–59*). For this purpose, frameworks such as hidden variable models or other recent dynamical models for social networks could be employed (*60–62*). Fourth, other assumptions may be instead optimistic, regarding the probability of detection of index cases, and compliance to isolation, for example. Few data are available to inform these parameters that may also vary over time (depending on the epidemic context and increased population fatigue) and across countries (depending on cultural aspects and regulations in place). While we explored a range of parameter values, more detailed information will be needed to contextualize our approach to a specific epidemic situation in a given country.

## Material and Methods

### Synthetic population

The model simulates the population of Metropolitan France representing individual inhabitants. This approach is similar to studies done previously e.g. for Italy(*27*) and for USA(*28*). The French synthetic population is based on the National Institute of Statistics and Economic Studies (INSEE) censuses. Individuals were assigned to municipalities according to the administrative borders. The number of households and the age structure of their inhabitants, sizes of schools and workplaces, fluxes of commuters between municipalities also followed the distribution of these statistics found in the INSEE data. Population size was kept constant through a simulation as we aimed at simulating one season of the epidemic.

To generate the population, we defined several statistics derived from INSEE publicly available data:

- The list of municipalities (“les communes de France”) of Metropolitan France (2015) with each municipality described by its INSEE code, population size, number of schools of six different levels (from kindergarten to university), number of workplaces in given size categories (0-9, 10-49, 50-99, 100-499, 500-999 and over 1000 employees) (Populations légales 2017, INSEE, https://www.insee.fr/fr/statistiques/4265429?sommaire=4265511).
- Statistics regarding the percentage of people in given age groups enrolled in each of six school levels, employed and unemployed (Bilan démographique 2010, INSEE, https://www.insee.fr/fr/statistiques/1280950).
- The age pyramid for France as the population fractions of individuals of a given age (Bilan démographique 2010, INSEE, https://www.insee.fr/fr/statistiques/1280950).
- The number of people commuting to work between each pair of municipalities (Mobilités professionnelles en 2016: déplacements domicile - lieu de travail, INSEE, https://www.insee.fr/fr/statistiques/4171554).
- The number of people commuting to school between each pair of municipalities (Mobilités scolaires en 2015: déplacements domicile - lieu d’études, INSEE, https://www.insee.fr/fr/statistiques/3566470).
- The probability distributions of sizes of households in France (Couples - Familles - Ménages en 2010. INSEE, https://www.insee.fr/fr/statistiques/2044286/?geo=COM-34150.)
- The probability of individuals belonging to a particular age class, given their role in the household: child of a couple, child of a single adult, adult in a couple without children, adult in a couple with children (Couples - Familles - Ménages en 2010. INSEE, https://www.insee.fr/fr/statistiques/2044286/?geo=COM-34150).

With the above statistics, the synthetic population was generated in the following steps:

1. Initialization of all the municipalities with an appropriate number of schools of each type and workplaces of given sizes.
2. Creation of schools in each municipality according to given statistics.
3. Creation of workplaces in each municipality according to given statistics.
4. Definition of the commuter fluxes between municipalities.

Each municipality has a defined number of inhabitants and individuals are created (one by one) until this number is reached. Each individual was assigned an age, a school or a workplace (or is assigned to stay at home) according to probability distributions derived from the data mentioned above.

The numbers of households within each municipality were not defined explicitly, but depended on the number of individuals. The municipal population size and statistics regarding family demographics constrain the number of households. Additional details on the algorithm for the population reconstruction are provided in the Supplementary Material.

### Face-to-face contact network

The synthetic population encodes information on the school, workplace, household and community each individual belongs to. We used this information to extract a dynamic network representing daily face-to-face contacts. We parametrized this network based on contacts’ statistics for the French population (*33*).

First, we generated a time aggregated network representing all contacts that can potentially occur – we will call this the *acquaintance network*, with some abuse of language since it includes also sporadic contacts. Second, to each contact we assigned a daily rate of activation. Then, in the course of the simulation we sampled contacts each day based on their rate.

The acquaintance network has five distinct layers representing contacts in household (layer *H*), workplace (layer *W*), school (layer *S*), community (layer *C*) and transports (layer *T*). The household layer is formed by a collection of complete networks linking individuals in the same household. The *W, S, C*, and *T* layers are formed by collections of Erdős– Rényi networks generated in each location *i*, with average degree *X*_*i*_ A location can be a workplace (*W* layer), a school (*S* layer) and a municipality (*C* and *T* layers). *X*_*i*_ is extracted at random for each place and depends on the type and size of the location. In particular, when the size of a location is small we assume that each individual enters in contact with all the others frequenting the same place. As the size increases the number of contacts saturates.

Once the acquaintance network was built a daily activation rate *x* was assigned to each link according to a cumulative distribution that depends on the layer *s*. For simplicity we assumed this distribution to be the same for *s* = *W, S, C*, while we allowed it to be different in household (where contacts are more frequent) and in transports (where contacts are sporadic).

Parameters were tuned based on average daily number of contacts, proportion of contacts by setting, and contact frequency as provided in (*33*) (Figure 1 C D). Additional details on the network reconstruction and parametrization are provided in the Supplementary Material.

### Transmission model

We defined a minimal model of COVID-19 spread in the general population that accounts for two levels of symptoms: none to mild (subclinical cases, *I*_*sc*._), and moderate to severe (clinical cases, *I*_*c*_). We assumed that clinical cases stay at home after developing symptoms. Susceptible individuals, if in contact with infectious ones, may get infected and enter the exposed compartment (*E*). After an average latency period *ϵ*^−1^ they become infectious, developing a subclinical infection with probability 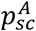 and a clinical infection otherwise. From *E*, before entering in either *I*_*sc*._ or *I*_*c*_, individuals enter first a prodromal phase (either *I*_*p,sc*,_ Or *I*_*p,c*,._), that lasts on average 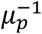 days and where individuals do not show any sign of illness, despite being already infectious. Contact-tracing, population-screening and modelling studies provide evidence that infectivity is related to the level of symptoms, with less severely hit individuals being also less infectious (*11, 43*). Therefore, we assumed that subclinical cases, *I*_*p,sc*,._ and *I*_*sc*._ have a reduced transmissibility compared to *I*_*p,c*,._ and *I*_*c*_. This is modulated by the scaling factor *β*_*I*_. We neglected hospitalization and death and assumed that with rate *μ* infected individuals become recovered.

The impact of COVID-19 is heterogeneous across age groups (*9–13*). This may be driven by differences in susceptibility (*9*), differences in clinical manifestation (*11, 13*) or both (*10*). We considered here both effects in agreement with recent modelling estimates (*10*). Susceptibility by age, *σ*_*A*_, was parametrized from (*9*), while clinical manifestation, 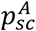, was parametrized from a large-scale descriptive study of the COVID19 outbreak in Italy (*13*).

Transition rates are summarized in Figure 2 B, and parameters and their values are listed in Table 1. The incubation period was estimated to be around 5.2 days from an early analysis of 425 patients in Wuhan (*34*). COVID-19 transmission potential varies across settings, populations and social contexts (*14–16*). In particular, indoor places were found to increase the odds of contagion 18.7 times compared to an open-air environment (*16*). In our model we assumed that all contacts at work, school and transport occur indoor and have the same transmission risk (*ω*_*S*_). In the contact survey of Béraud et al. (*33*), 46% of contacts in the community were occurring outdoors. Combining this information with the 18.7 indoor vs. outdoor risk ratio leads to a 60% relative risk of community contacts with respect to workplace/school/transport contacts. Contacts within households are generally associated to a higher risk with respect to other settings, because they last longer and there is a higher risk of environmental transmission. We assumed that the transmission risk associated with them is twice the one in workplace/school/transport. For the basic reproductive ratio of COVID-19 we took *R*_0_~3.1 (*1, 36*). We also explored lower levels of transmission potential, i.e. reproductive ratios *R* down to 1.5, to effectively account for behavioral changes and adoption of barrier measures. Our definition of *R* does not integrate population immunity. We explicitly indicate the initial level of population immunity to disentangle the relative role of the two quantities.

### Modelling contact tracing

#### Self-isolation and isolation of household contacts

Self-isolation and isolation of household contacts was modelled according to following rules:

- As an individual shows clinical symptoms, s/he is detected with probability *p*_*d,c*._ (baseline value 50%, additional explored values 30% and 80%). If detected, case confirmation, isolation and contacts’ isolation occur with rate *r*_*H*,._ = _0_.9 upon symptoms onset.
- Subclinical individuals are also detected with probability *p*_*d,sc*._ (baseline value 5%, additional explored values 25% and 45%), and rate *r*_*d,sc*._ = 0.5.
- The individual’s family members are isolated with probability *p*_*c,h*_ = 0.9 (0.5 and 0.7 were also explored).
- We assume that contacts are tested and the follow up guarantees that all individuals who got infected prior to isolation are detected. Thus, contacts that are negative (either susceptible or recovered at the time of isolation) terminate their isolation after 7 days. The index-case and the positive contacts are isolated for 14 days. Contacts with no clinical symptoms have a daily probability *p*_*drop*_ =0.02 to drop-out from isolation.
- For both the case and the contacts, isolation is implemented by assuming no contacts outside the household and contacts within a household having an associated transmission risk (i.e. the weight *ω*_*H*_) reduced by a factor *ι* = 0.5.

#### Digital contact tracing

We assumed that contact tracing is adopted in combination with self-isolation and isolation of household members. Therefore, we added the following rules to the ones outlined above:

- At the beginning of the simulation, a smartphone is assigned to individuals with probability 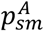, based on the statistics of smartphone penetration (0% for [0,11], 86% for [12,17], 98% for [18,24], 95% for [25,39], 80% for [40,59], 62% for [60,69], 44% for 70+) (*29*).
- Each individual with a smartphone has a probability *p*_*S*_ to download the app (we explored values between 0 and 0.9).
- Only contacts occurring between individuals with a smartphone and the app are traced.
- If the individual owns a smartphone and downloaded the app the contacts that s/he has traced in the period since *D* = 7 days before his/her detection are isolated with probability *p*_*c,a*_ = 0.9 (0.5 and 0.7 were also tested).
- We assume contacts are tested and the follow up guarantees that all individuals who got infected prior to isolation are detected. Thus, contacts that are negative (either susceptible or recovered at the time of isolation) terminate their isolation after 7 days. The index-case and the positive contacts are isolated for 14 days. Contacts with no clinical symptoms have a daily probability *p*_*drop*_ = 0.02 to drop out from isolation.
- For both the case and the contacts, isolation is implemented by assuming no contacts outside the household and contacts within a household having an associated transmission risk (i.e. the weight *ω*_*H*_) reduced by a factor *ι* = 0.5.

## Data Availability

All data needed to evaluate the conclusions in the paper are present in the paper or available from public sources cited on the paper.

## Author contributions

CP conceived the study; PYB, VC, AB, CP designed the study and interpreted the results; CP, wrote the original draft; PB, LB, FP implemented the synthetic population model; JAML, BAG, implemented the epidemic model; All authors edited the manuscript and approved its final version.

## Data and materials availability

Figures and Tables

## Supplementary Material of

### Additional methods

#### Algorithms for the generation of the synthetic population

The generation of the synthetic population is a stochastic process resulting in a contact network being slightly different each time it is generated. Thanks to this mechanism, we can account for some of the uncertainty concerning the input data. It also allows for population scaling to reduce the population by respecting its composition and spatial distribution, thus increasing computational efficiency. Specifically, the scaling decreases the number of individuals in municipalities and the fluxes of commuters between them, but it does not impact the number of municipalities nor the number of schools and workplaces. The smaller population has a smaller number of households, but it maintains the statistics regarding family size and age structure given by the INSEE data.

### Households

Census data on age structure and household type and size are used to randomly assign age and locate individuals in households. Five different types of household are considered: single person, single with children, couple without children, couple with children, other household groups; some of the household types may also contain an additional adult member (usually an elderly person or a relative: if the number of additional adults is greater than one, the household falls in the “other” category). For each municipality *m* with population size *pop*_*m*_, we generate new households until the size of the virtual population of the municipality *vir*_*m*_reaches the real size of population *pop*_*m*_. For each household, we determine its type, its size and the presence of an additional member (if the household type is not single-person or couple without children, and in the case of a household with children if the size is greater than the number of adults plus one): then, according to the role of each individual (adult, child or other) we randomly extract his/her age, with some additional conditions:

C1: the age of any child is between 15 and 45 years less than that of the youngest parent;

C2: spouses’ ages differ by no more than 15 years.

The detailed procedure is summarized in Algorithm 1.

#### Algorithm S1: Creation of households in municipalities

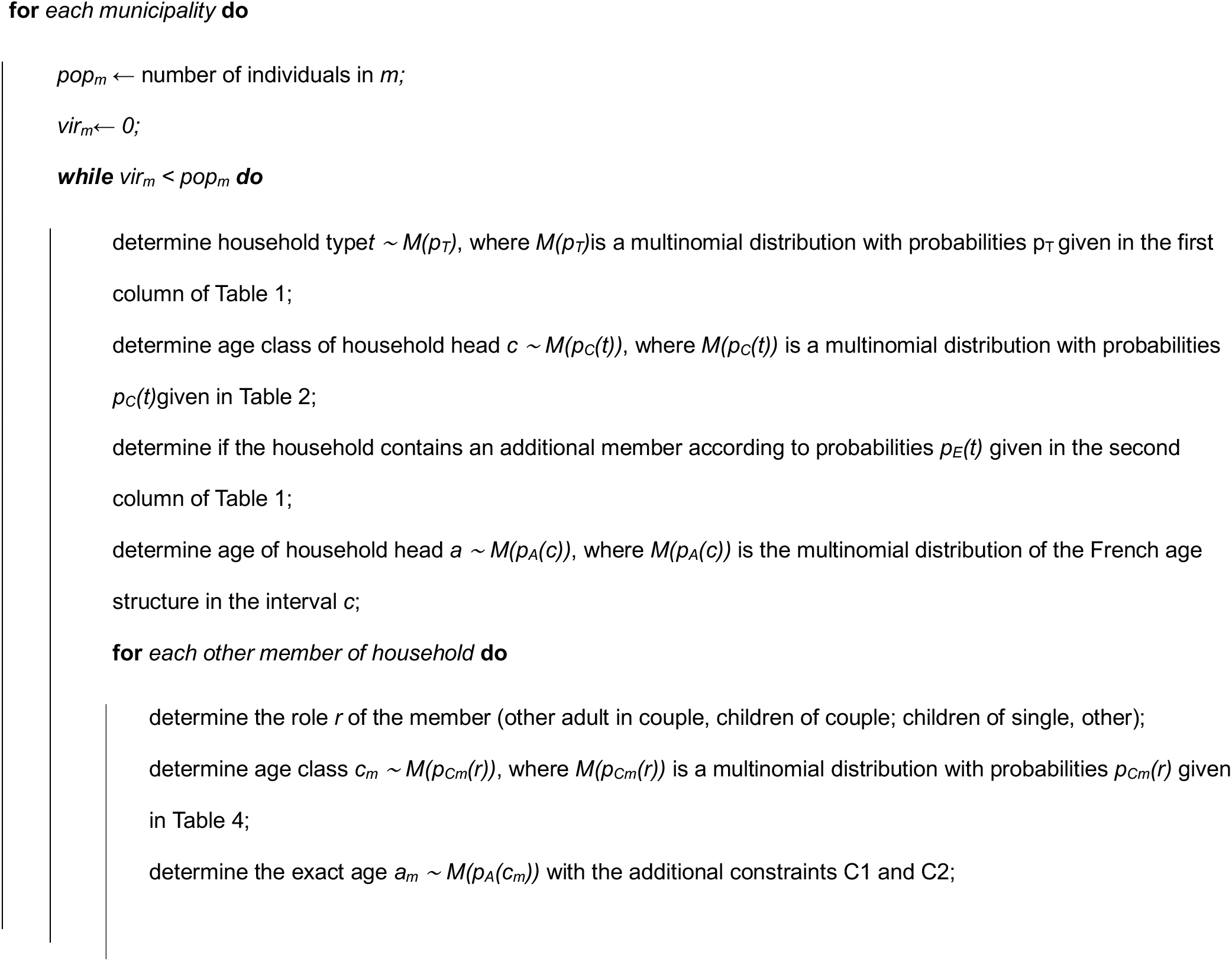
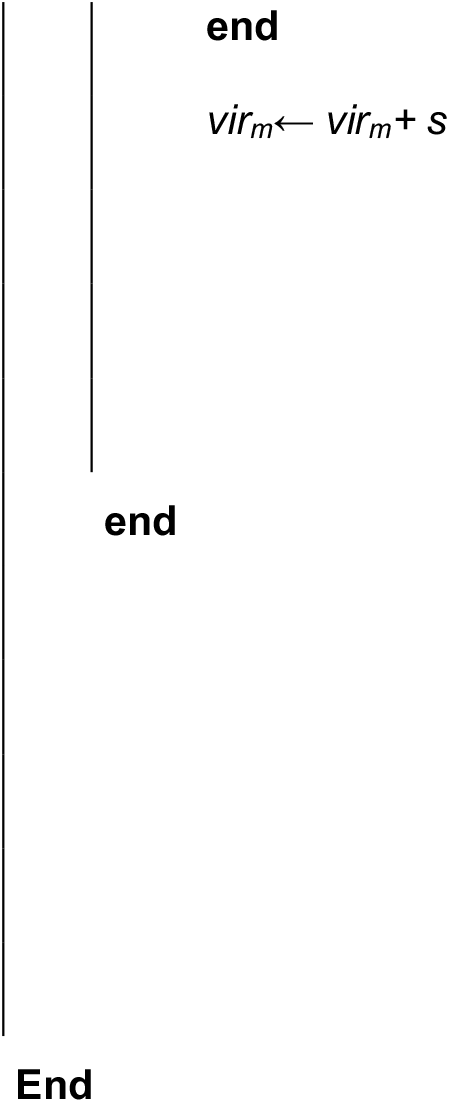

**Table S1.** For each household type, the fist column shows its frequency; the second column the probability that the household type contains an additional member; the third column the frequency of individuals in each household type

| Type of household | Frequency | Fr. of add. member | Fr. of individuals |
| --- | --- | --- | --- |
| Single without children | 0.338 | 0.0 | 0.149 |
| Couple without children | 0.268 | 0.0204 | 0.243 |
| Couple with children | 0.277 | 0.0096 | 0.475 |
| Single with children | 0.09 | 0.0123 | 0.106 |
| Other | 0.027 | 0.0 | 0.027 |

**Table S2.** For each household type, the frequency of the age class of the household head

| Type of household | 0 - 14 | 15 - 19 | 20 - 24 | 25 - 39 | 40 - 54 | 55 - 64 | 65 - 79 | 80 - 100 |
| --- | --- | --- | --- | --- | --- | --- | --- | --- |
| Single without children | 0.0 | 0.019 | 0.075 | 0.191 | 0.186 | 0.162 | 0.212 | 0.155 |
| Couple without children | 0.0 | 0.029 | 0.132 | 0.263 | 0.264 | 0.133 | 0.094 | 0.085 |
| Couple with children | 0.0 | 0.002 | 0.023 | 0.271 | 0.47 | 0.116 | 0.073 | 0.045 |
| Single with children | 0.0 | 0.001 | 0.021 | 0.251 | 0.313 | 0.197 | 0.17 | 0.047 |
| Other | 0.0 | 0.008 | 0.042 | 0.233 | 0.283 | 0.176 | 0.173 | 0.085 |

**Table S3.**
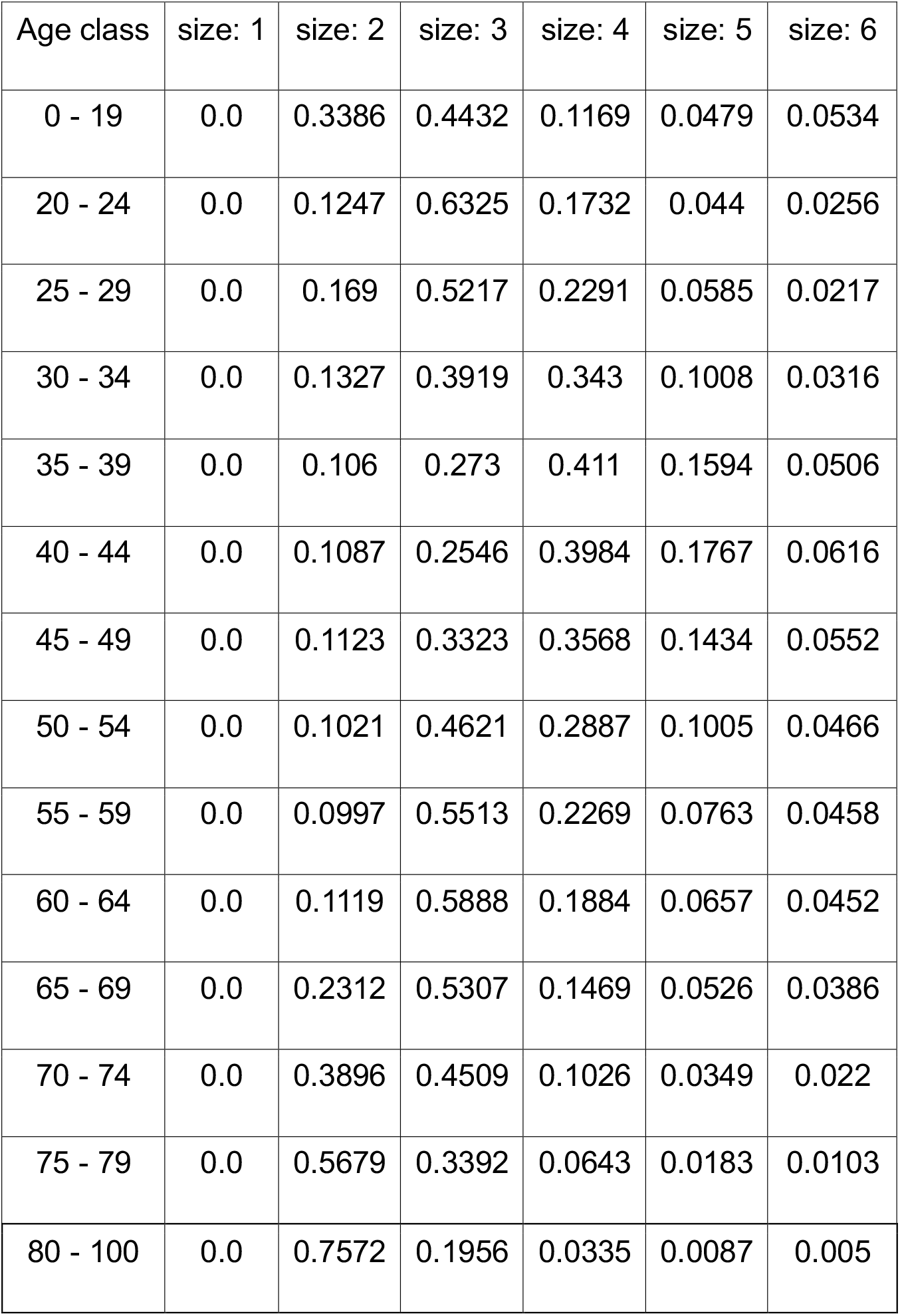
Probability distributions of the size of the household (except singles and couples without children, having size 1 and 2, respectively) for each age class of the household head. Rows are the age class of the household head, while columns are the size of the household

| Age class | size: 1 | size: 2 | size: 3 | size: 4 | size: 5 | size: 6 |
| --- | --- | --- | --- | --- | --- | --- |
| 0 - 19 | 0.0 | 0.3386 | 0.4432 | 0.1169 | 0.0479 | 0.0534 |
| 20 - 24 | 0.0 | 0.1247 | 0.6325 | 0.1732 | 0.044 | 0.0256 |
| 25 - 29 | 0.0 | 0.169 | 0.5217 | 0.2291 | 0.0585 | 0.0217 |
| 30 - 34 | 0.0 | 0.1327 | 0.3919 | 0.343 | 0.1008 | 0.0316 |
| 35 - 39 | 0.0 | 0.106 | 0.273 | 0.411 | 0.1594 | 0.0506 |
| 40 - 44 | 0.0 | 0.1087 | 0.2546 | 0.3984 | 0.1767 | 0.0616 |
| 45 - 49 | 0.0 | 0.1123 | 0.3323 | 0.3568 | 0.1434 | 0.0552 |
| 50 - 54 | 0.0 | 0.1021 | 0.4621 | 0.2887 | 0.1005 | 0.0466 |
| 55 - 59 | 0.0 | 0.0997 | 0.5513 | 0.2269 | 0.0763 | 0.0458 |
| 60 - 64 | 0.0 | 0.1119 | 0.5888 | 0.1884 | 0.0657 | 0.0452 |
| 65 - 69 | 0.0 | 0.2312 | 0.5307 | 0.1469 | 0.0526 | 0.0386 |
| 70 - 74 | 0.0 | 0.3896 | 0.4509 | 0.1026 | 0.0349 | 0.022 |
| 75 - 79 | 0.0 | 0.5679 | 0.3392 | 0.0643 | 0.0183 | 0.0103 |
| 80 - 100 | 0.0 | 0.7572 | 0.1956 | 0.0335 | 0.0087 | 0.005 |

**Table S4.** Probability of age class of individuals depending on his role in the household, excluding the household head

| Age class | Child of couple | Child of a single | Adult of couple with children | Adult of couple without children | Other |
| --- | --- | --- | --- | --- | --- |
| 0 - 4 | 0.2384 | 0.1288 | 0.0 | 0.0 | 0.0161 |
| 5 - 9 | 0.2264 | 0.181 | 0.0 | 0.0 | 0.0176 |
| 10 - 14 | 0.212 | 0.2125 | 0.0 | 0.0 | 0.0226 |
| 15 - 19 | 0.1777 | 0.2131 | 0.0035 | 0.0008 | 0.0825 |
| 20 - 24 | 0.0886 | 0.1144 | 0.0445 | 0.0164 | 0.1587 |
| 25 - 29 | 0.0305 | 0.0448 | 0.071 | 0.0718 | 0.1051 |
| 30 - 34 | 0.0105 | 0.0204 | 0.0414 | 0.1392 | 0.066 |
| 35 - 39 | 0.0064 | 0.0182 | 0.0244 | 0.1848 | 0.0576 |
| 40 - 44 | 0.0045 | 0.0181 | 0.0224 | 0.1873 | 0.0577 |
| 45 - 49 | 0.0028 | 0.0172 | 0.0434 | 0.1642 | 0.0578 |
| 50 - 54 | 0.0014 | 0.0139 | 0.0902 | 0.111 | 0.057 |
| 55 - 59 | 0.0006 | 0.0101 | 0.1379 | 0.0621 | 0.057 |
| 60 - 64 | 0.0002 | 0.0053 | 0.1537 | 0.0306 | 0.0506 |
| 65 - 69 | 0.0 | 0.0016 | 0.1126 | 0.0135 | 0.0364 |
| 70 - 74 | 0.0 | 0.0005 | 0.0993 | 0.0086 | 0.0356 |
| 75 - 79 | 0.0 | 0.0001 | 0.0812 | 0.0056 | 0.0382 |
| 80 - 100 | 0.0 | 0.0 | 0.0745 | 0.0041 | 0.0835 |

### Employment

School and industry census data are used to randomly assign an employment category to each individual on the basis of age: the probabilities are reported in Table S5. The table assumes that all the children attend school from elementary to high school, while the attendance of universities is taken from census data. Census data contains the number of individuals commuting from one municipality to another one, specifying if they are students or workers, and the same for the number of individuals that study or work in the same municipality where they live. Such information is used to randomly assign students and workers to a municipality of work, which may be different from the one where they live, and then to a random school building or workplace inside this municipality. Students are assigned to a random school building, while workers are assigned to a random workplace type (5 types, depending on the workplace size, i.e., the number of employees) and then to a random workplace building. Students are not grouped in classes.

**Table S5.** Probability of workplace kind by age: individuals in the last column stay at home

| Age<br>class | creche | maternel | elementaire | college | lycee | universite | work | neet |
| --- | --- | --- | --- | --- | --- | --- | --- | --- |
| 0 - 3 | 0.6 | - | - | - | - | - | - | 0.4 |
| 4 - 6 | - | 1.0 | - | - | - | - | - | - |
| 7 - 11 | - | - | 1.0 | - | - | - | - | - |
| 12 - 15 | - | - | - | 1.0 | - | - | - | - |
| 16 - 18 | - | - | - | - | 1.0 | - | - | - |
| 19 - 21 | - | - | - | - | - | 0.365 | 0.448 | 0.187 |
| 22 - 23 | - | - | - | - | - | 0.332 | 0.471 | 0.197 |
| 24 - 26 | - | - | - | - | - | 0.026 | 0.687 | 0.287 |
| 27 - 62 | - | - | - | - | - | - | 0.705 | 0.295 |
| 63 - 100 | - | - | - | - | - | - | - | 1.0 |

## Face-to-face contact network

### Acquaintance network

For each place, *i* – a workplace (*W* layer), a school (*S* layer) and a municipality (*C* and *T* layers) – we build an Erdős–Rényi network, with average degree *X*_*i*_. The latter is a stochastic variable and depends on place layer, *s*_*i*_, and size, *n*_*i*_. We draw it from a gamma distribution with average 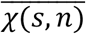 and coefficient of variation *CV*.

We expect that when the size of a place is small each individual enters in contact with everybody. As the size increases the number of contacts saturates. We model this by assuming 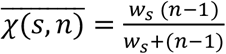. The function approaches (*n* − 1) for small *n* and saturates to *wS* as *n* increases (Figure S1).

For each setting the parameter *w*_*s*_ is tuned to reproduce the overall proportion of contacts occurring in the layer. *CV* rules the level of heterogeneity among places of the same kind and size. For simplicity we assume it to be the same for all settings. Additional details on the parametrization are provided below.

**Figure S1.**
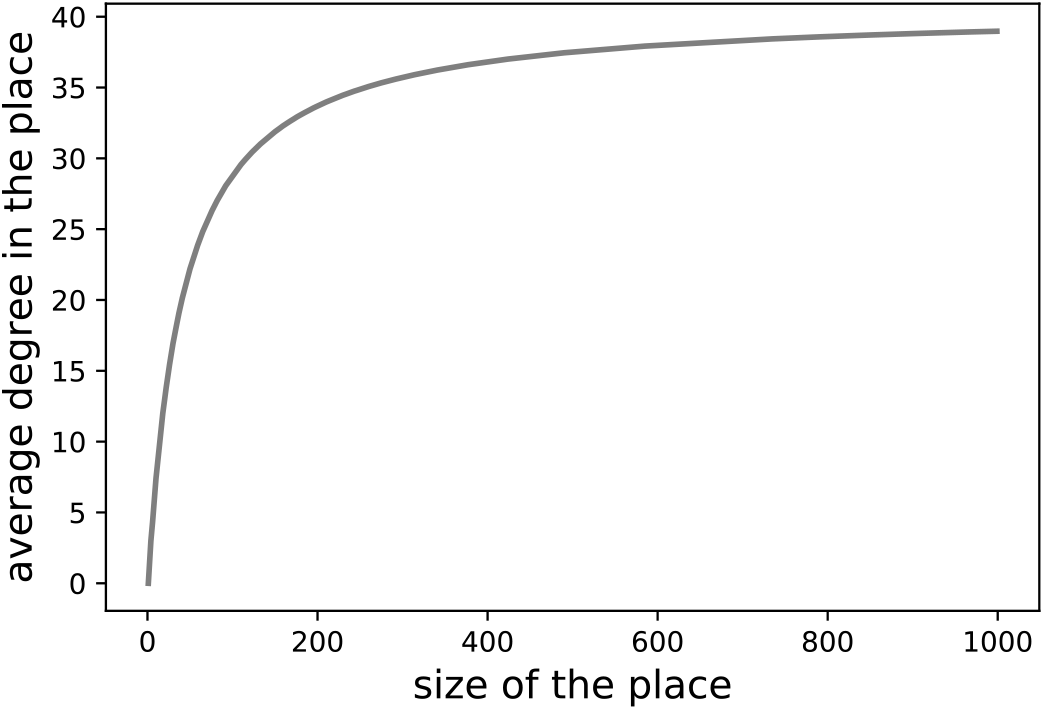
Average degree of the acquaintance network 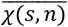 as a function of the size of the place. As the size goes to infinity the degree saturates to *w*_*s*_ that depends on the setting. Here we show as an example the curve for the workplace (*w*_*w*_ = 41.8). The other parameters estimated are *w*_*s*_ = 18,23, *w*_*c*_ = 4,3, *w*_*T*_ = 2_0_,9.

### Daily contact network

Once the acquaintance network is built a daily activation rate *x* is assigned to each link according to a cumulative distribution *F*_*S*_(*x*) that depends on the layer *s*. We model *F*_*s*_(*x*) with a sigmoid function with two parameters, *A*_*S*_ and *B*_*S*_. For simplicity we assume *F*_*s*_(*x*) to be the same for *s* = *W, S, C*, while we allow it to be different in households (where contacts are more frequent) and in transports (where contacts are sporadic). On average, a fraction ⟨*x*⟩_*s*_ of the links of the acquaintance network is active each day. By indicating with *K*_*s*_ and *K*_*s*_ the average daily number of contacts networks, respectively, we have 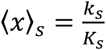

### Parametrization

We tuned parameters *w*_*s*_, *A*_*s*_, *B*_*s*_ and *CV* to reproduce the contact statistics in (*33*), namely:

- the average daily number of contacts is 10;
- the contact distribution is skewed with mode 3;
- being *K*_*s*_ the average daily number of contacts in setting *s* and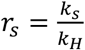 the survey reports *r*_*W*_ = 3.17, *r*_*S*_ = 1.55, *r*_*c*_ = 1.86, and *r*_*T*_ = _0_.23.
- 35% of the registered contacts were with people met every day, while the rest with people met less frequently.

Specifically, these data provide the following constraints:

- Combining point 1) and 2) above we get *K*_*H*_(1 + *r*_*W*_ + *r*_*S*_ + *r*_*c*_) = 1_0_, meaning *K*_*H*_ = 1.28. The household statistics used for our synthetic population reconstruction yields *K*_P_ = 1.97. This implies ⟨*x*⟩_*H*_ = _0_.65.
- We assume that the daily contact network has 35% of links with activity rate >0.95. In order to do so we must account for the fact that being f_*S*_(*x*) the distribution of activation rate values assigned to links of the acquaintance network (i.e. the probability density function associated to *F*_*S*_(*x*)), the distribution of links sampled in the daily network is biased toward higher rates, i.e. it is given by 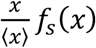

In addition to these, we assume uncommon but still possible to meet more than once people in transports within a time frame of one or few months. Thus, we assume an activation rate for links in the transport layer as high as a few percent or lower. Based on these constraints we design the frequency distribution as in Figure S2.

Once *F*_*s*_(*x*) is parametrized we tune the parameters *w*_*s*_ of the acquaintance network to reproduce the proportion *r*_*s*_ of daily contacts in different settings. We then fix *CV* = _0_.2 to reproduce the mode of the distribution. The main properties of the network (contact distribution, link activation frequency, and age contact matrix) are shown in Figure 1 of the main paper. Other features are summaries in Table S1.

**Figure S2.**
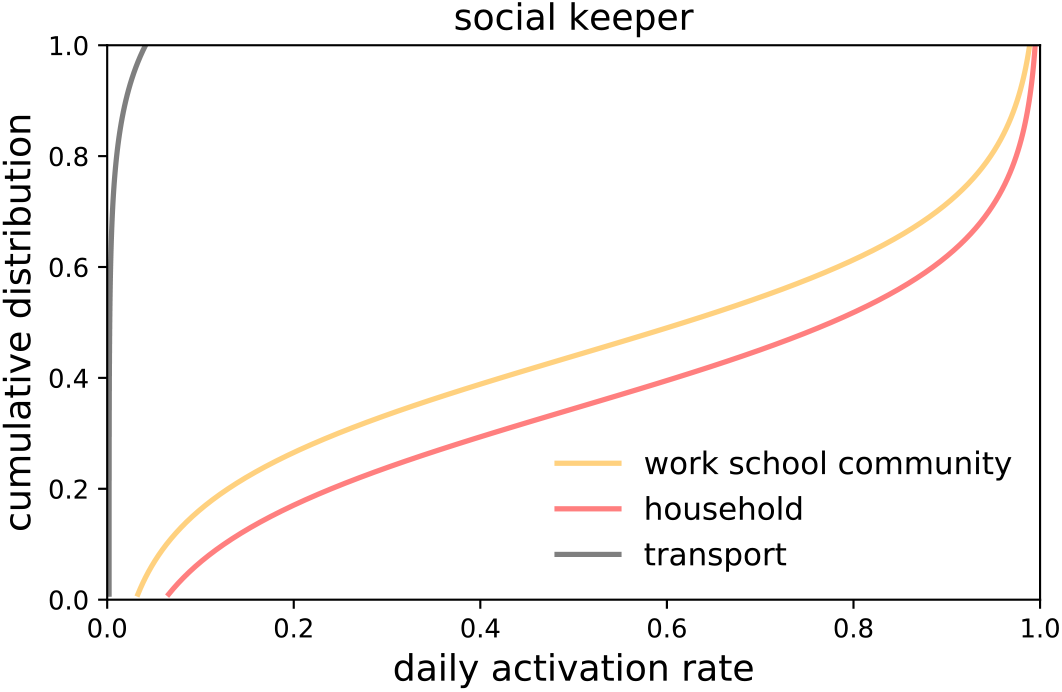
Cumulative distribution of the daily activation rate of contacts. We model it with the function *F*(*x*) = *A* tanh^−^(1 − 2*x*) + *B*. Parameters are the following: *A* = 0.25 for all settings; *B*_*H*_ = 0.65,*B*_*T*_ = −0.4_0_, *B*_*S*_ = 0.56 (*s* = *S, W, C*).

**Table S6.** Main network features.

| Parameter | Description | Value |
| --- | --- | --- |
| $\langle x \rangle_s, s = S, W, C$ | Average activation rate of contacts for the school, workplace, community layer | 0.56 |
| $\langle x \rangle_H$ | Average activation rate of contacts for the household layer | 0.65 |
| $\langle x \rangle_T$ | Average frequency of contacts for the transport layer | 0.007 |
| $k_H (K_H)$ | Average degree of the daily network (acquaintance network) in a household | 1.79 (2.31) |
| $k_W (K_W)$ | Average degree of the daily network (acquaintance network) in a workplace | 12.9 (22) |
| $k_S (K_S)$ | Average degree of the daily network (acquaintance network) in a school | 7.9 (14) |
| $k_C (K_C)$ | Average degree of the daily network (acquaintance network) in a community | 2.6 (4.3) |
| $k_T (K_T)$ | Average degree of the daily network (acquaintance network) in transport | 1.1 (21) |

### Details of the numerical simulations

Simulations are discrete-time and stochastic. At each time step, corresponding to one day, three processes occur: *(i)* the contact network is sampled according to the activation rate of each link; *(ii)* for each node, the infectious status is updated; and *(iii)* cases and contacts are isolated, or get out from isolation.

The transmission process is modelled through the links of the multi-layer network as follows. At each time step, a susceptible node *i* gets infected with the following probability

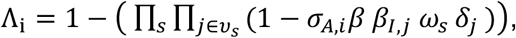

where *j* is a node belonging to the neighborhood *v*_*s*_ of *i* on layer *s*, and *δ*_*j*_ is 1 if *j* belongs to any infectious stage (*I*_*p,sc*._, *I*_*p,c*_, *I*_*sc*._, *I*_*c*._) and 0 otherwise. Links of layer *s* have weight *ω*_*s*_, that represent the average level of risk associated to contacts occurring in the setting *s*. We assume that individuals in the *I*_*c*_ state stay at home due to illness and can therefore transmit the disease only through the links of the household layer.

Simulations are run on the synthetic municipality of Strasbourg. The population is reduced by a factor 3 to shorten simulation time to feasible levels, yielding a population of 92,423 individuals.

A single-run simulation is executed with no modelled intervention, until the desired immunity level is reached. This guarantees that immune individuals are realistically clustered in space. Then the simulations of the epidemic with contact tracing are started, considering 15 individuals initially infected randomly assigned in the population.

Quantities shown in Figures 2, 3, 4, and 5 of the main paper are computed by averaging results over different stochastic realizations. We run 100 stochastic simulations. Increased statistics (300 runs) was necessary to accurately compare the relative reduction in incidence obtained with low app adoption levels – i.e. household isolation only, app adoption 6% and app adoption 19%.

We vary COVID-19 transmission potential by tuning the daily transmission rate per contact *β*. The reproductive number *R* is computed numerically as the average number of infections each infected individual generates throughout its infectious period. To do so, population immunity at the beginning of the simulation is set to 0 and *R* is computed considering the infections generated by individuals who get infected the first two time-steps of the simulations to guarantee that the whole population is susceptible. We find that *β* = 0.1, 0.125,0.15, …, 0.25 corresponds to the following *R* values: 1.47 95% CI [0.0, 4.86], 1.75 95% CI [0.0, 5.78], 2.05 95% CI [0.0, 5.31], 2.25 95% CI [0.0, 5.31], 2.61 95% CI [0.0, 6.13], 2.95 95% CI [0.06, 7.29], 3.09 95% CI [0.68, 8.11]. We also computed numerically the generation time from the infector-infected pairs.

## Additional results

### Uncontrolled epidemic

**Figure S3.**
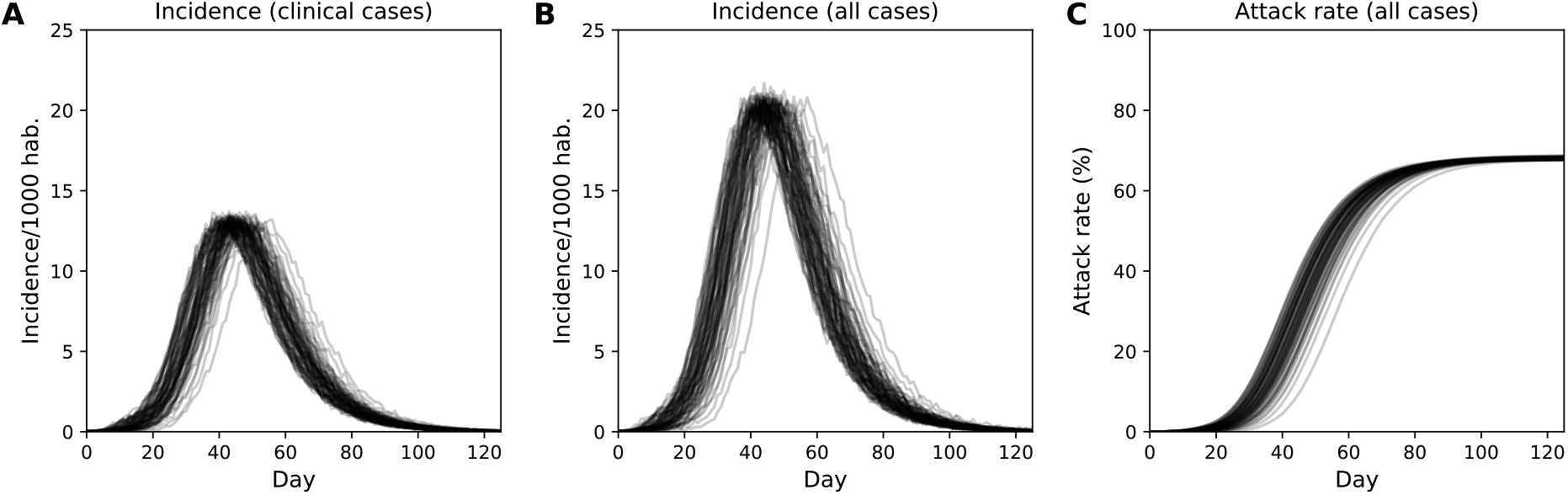
Epidemic in an uncontrolled scenario. **A** Incidence of clinical cases. **B** Incidence of all cases. **C** Attack rate. The bundle of curves shows 70 stochastic realizations. The epidemic is obtained with transmission rate *β* = 0.25 corresponding to *R*_/_ = 3.1.

### App adoption variable by age

The model accounted for age-varying smartphone penetration. However, we assumed that the probability of downloading the app, provided an individual owns a smartphone, is uniform and independent on age – uniform app adoption scenario, *U*. This hypothesis is simplified. Indeed, elderly people may be less inclined to use the app even when they own a smartphone. We also tested the extreme case scenario in which no individual in the 70+ age cohort adopts the app – non-uniform app adoption scenario, *NU*. We consider a 32% app coverage over the whole population, and we compared *U* and *NU* scenarios, assuming the same number of apps are downloaded in the two cases. Figure S4 shows the attack rate relative reduction 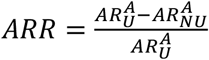 by age group, where 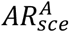 is the attack rate of the epidemic for the age group, *A*, and the scenario, *sce* = *U, NU*. We found that *ARR* is close to zero, meaning that 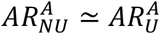, for all age groups (Figure S4). This means that distributing only to individuals younger than 70 years would not reduce the protection in the 70+ age group.

**Figure S4.**
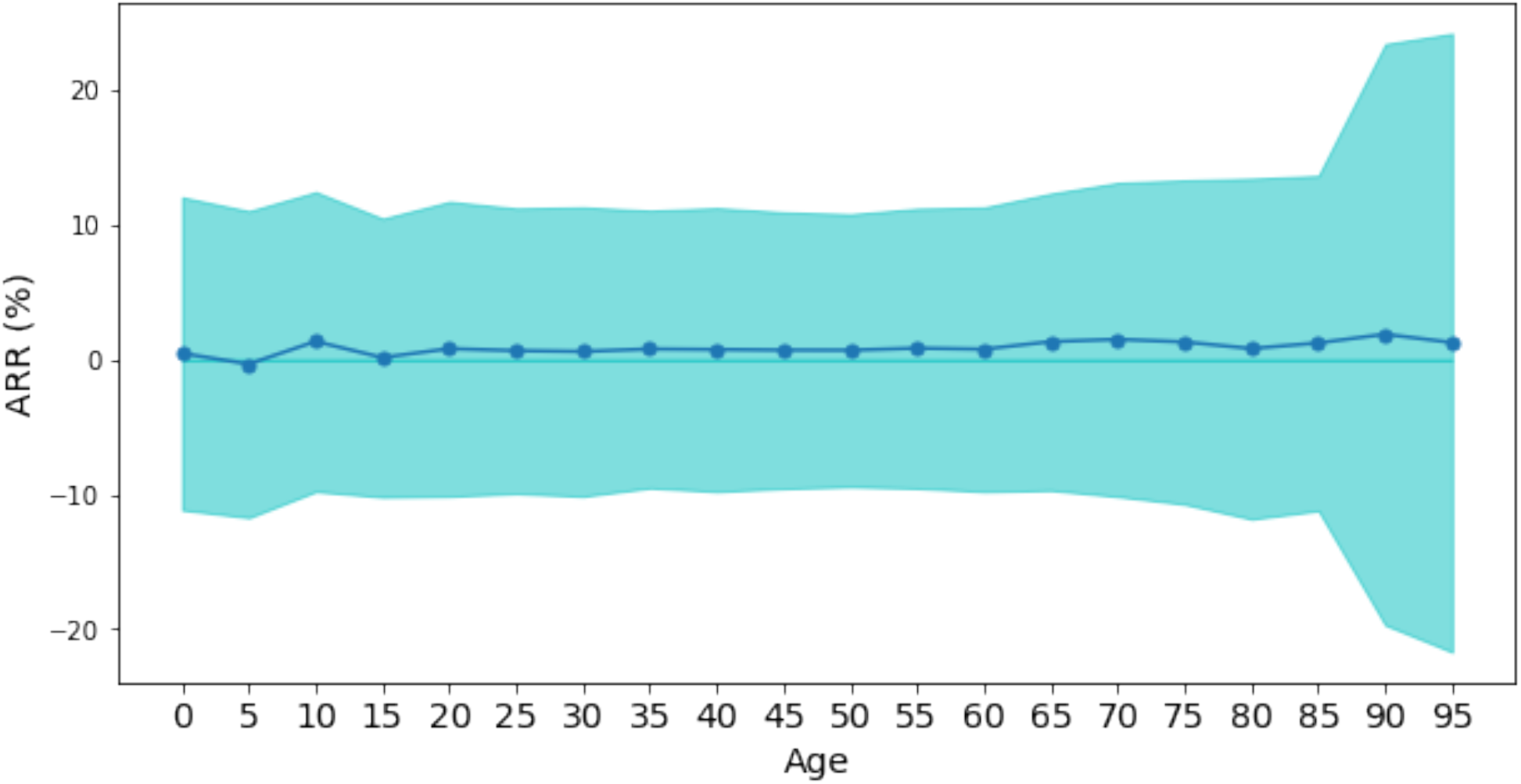
Comparison between the *U* and the *NU* contact tracing scenario. Here *R* = 2.6, Immunity is 10%, detection probability is 50% and app penetration is 32%. The line shows the average attack rate relative reduction and the shaded area is standard deviation.

